# Seasonal vaccine-induced immunity shows preserved cross-reactivity to H3N2 subclade K in adults

**DOI:** 10.64898/2026.02.18.26346502

**Authors:** Adria Wilson, Brian Lerman, Reima Ramsamooj, Jacob Mischka, Jordan Ehrenhaus, Ashley Aracena, Yusuf Figueroa, Keith Farrugia, Ana S. Gonzalez-Reiche, Jessica Nardulli, Zain Khalil, Charles Gleason, Eniko Hermann, Komal Srivastava, Emilia Mia Sordillo, Harm van Bakel, Anass Abbad, Florian Krammer, Viviana Simon

**Affiliations:** Department of Microbiology; Center for Vaccine Research and Pandemic Preparedness; Division of Infectious Diseases, Department of Medicine; Department of Pathology, Molecular and Cell Based Medicine; Department of Genetics and Genomic Sciences; The Global Health and Emerging Pathogen Institute; Department of Artificial Intelligence and Human Health; Icahn Genomics Institute; Icahn School of Medicine at Mount Sinai, New York; Ignaz Semmelweis Institute, Interuniversity Institute for Infection Research, Medical University of Vienna, Vienna, Austria; Ludwig Boltzmann Institute for Science Outreach and Pandemic Preparedness at the Medical University of Vienna, Vienna, Austria

**Keywords:** vaccine-induced immunity, influenza A H3N2 subclade K, antigenic drift, hemagglutination inhibition antibodies, neutralizing antibodies, and/or cross-reactive immunity

## Abstract

Influenza A subclade K viruses caused high infection rates in the 2025/2026 Northern Hemisphere season, raising concerns about antigenic drift and reduced vaccine effectiveness. We measured antibody responses in matched human pre- and post-vaccination sera against a vaccine-like as well as subclade K isolates. Pre-existing immunity to subclade K variants was noted with seasonal influenza vaccination boosting titers two-fold against subclade K and three-fold against the vaccine-like strain, indicating little antigenic drift. These data contrast ferret-based predictions of marked antigenic drift.

## Manuscript

### Antigenic drift of H3N2 subclade K in 2025-2026

Influenza A virus H3N2 strains are known to cause severe epidemics, particularly affecting older adults with high excess mortality and morbidity^1^. In the past six months^2^, a drifted H3N2 variant — initially termed J.2.4.1 and now commonly referred to as subclade K — emerged and spread rapidly, leading to an early onset of the influenza season in the Northern Hemisphere and record high numbers of influenza-like illnesses^3, 4^. H3N2 subclade K viruses possess several mutations in the antigenic sites of the hemagglutinin (HA) surface protein, which are associated with immune escape^2, 4, 5^. Studies using ferret sera raised against the current egg-based vaccine strain (A/Croatia/10136RV/ 2023) demonstrated more than 32-fold reductions in hemagglutination inhibition (HI) titers against subclade K viruses, while ferret sera from the cell-based vaccine strain (A/District of Columbia/27/2023) showed 8-to 16-fold reductions^4, 5^. These results indicate significant antigenic drift and raise concerns about vaccine effectiveness (VE), suggesting a potential mismatch between the H3N2 vaccine strains and circulating subclade K viruses^4^. However, initial VE measurements in the UK have shown good to moderate effectiveness^4, 6^.

To investigate these contradictory findings, we measured hemagglutination inhibition (HI) and virus neutralization titers in human sera collected before and after seasonal trivalent immunization with the 2025/26 influenza vaccine formulation. Using the Mount Sinai Pathogen Surveillance Program’s H3N2 genotypic tracking, we identified residual diagnostic nasal swabs suitable for culturing primary isolates representative of the cell-based H3N2 vaccine component (A/District of Columbia/27/2023) and three subclade K field isolates, which included viruses with and without additional substitutions in the antigenic sites of hemagglutinin compared to the clade K root (Fig. 1A; hemagglutinin structure visualized using the Chai-1 modeling tool^7^). To assess both pre-existing immunity and vaccine-induced responses, we selected paired sera collected before and after seasonal influenza vaccination from participants in ongoing IRB-approved longitudinal, observational studies. Sera were obtained from 100 adults (68% female, average age 31 years [range: 23–81]) who received either cell-based (N=83), recombinant (N=2), or egg-based (N=15) influenza vaccines (trivalent 2025/2026 formulation; see Table 1).

**Figure 1:**
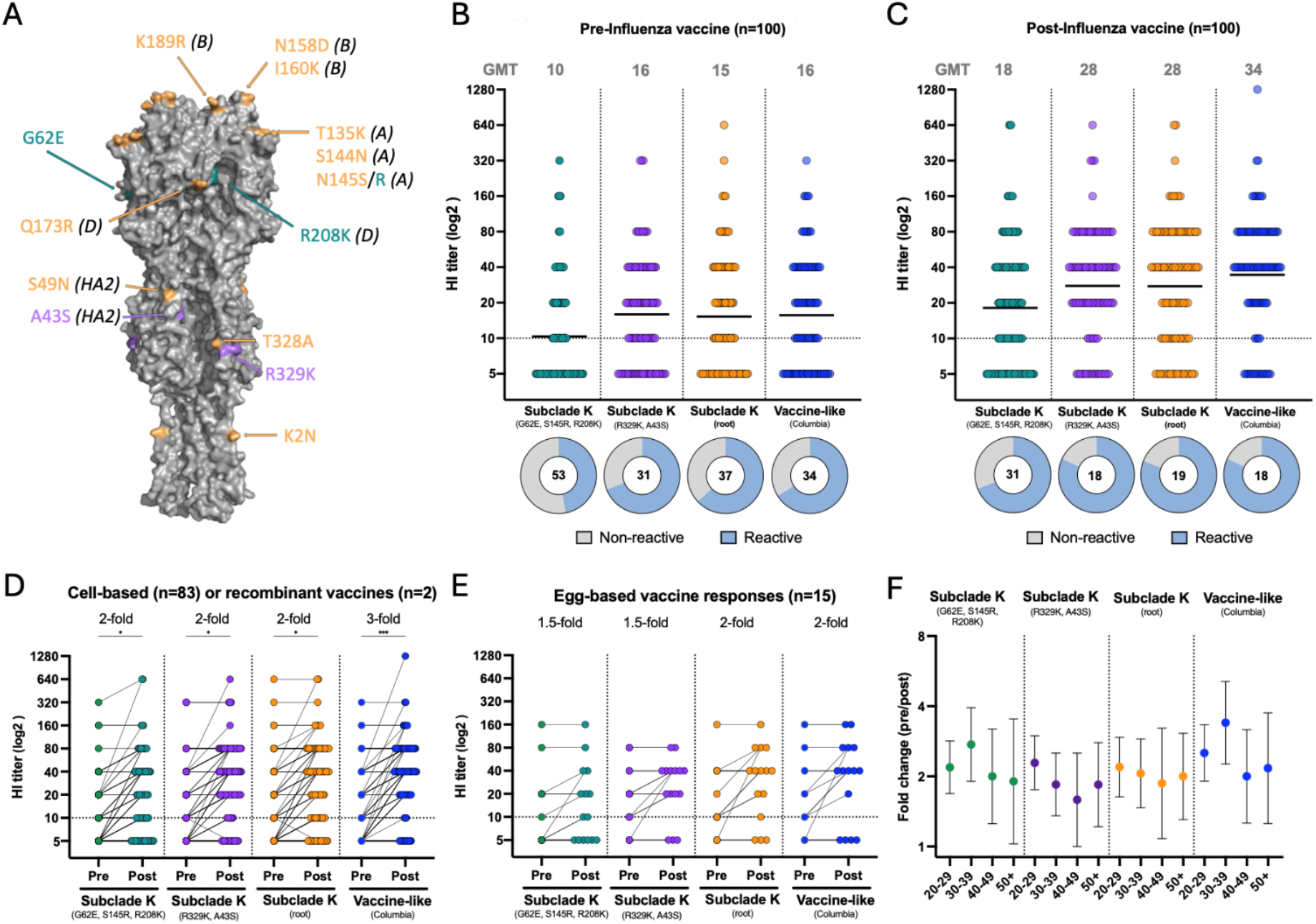
Hemagglutinin inhibiting antibody responses against H3N2 subclade K and vaccine like isolates before and after seasonal immunization. **A**: A model of the full-length H3 hemagglutinin from isolate A/New York City/PX23710 (subclade K root). The model is represented as HA0 and includes the transmembrane region. Amino acids that differ from the vaccine representative (A/New York City/PX12775/2024) are colored yellow, while the additional mutations of A/New York City/PX23641/2025 and A/New York City/ PV307069/2025 are shown in purple or green, respectively. Known antigenic escape sites (e.g., A, B, D) are listed in parentheses after the substitution. The model was prepared using Chai-1^7^ and visualized using PyMol. **B/C**: The pre- and post-influenza vaccine HI titers for 200 sera are shown for the four viruses tested. The donuts illustrate the proportion of reactive (HI titer: ≥10) and non-reactive sera with the number of sera with values below the limit of detection being shown for each virus and time point. **D**: HI titers measured in sera collected before and after vaccination with cell-based or recombinant influenza vaccine types are depicted. **E**: HI titers measured in sera collected before and after vaccination with egg-based influenza vaccine types are depicted. **F**: Geometric mean fold changes of the HI titers (post/pre, +/-SD) are shown for four different age groups (20-29 years: N=42; 30-39:N=27; 40-49: N=11; 50 and older: N=20).

**Table 1.**

| Vaccine Type | Cell-Based<br>(n=83) | Egg-Based<br>(n=15) | Recombinant<br>(n=2) | Total<br>(n=100) |
| --- | --- | --- | --- | --- |
| **Age at vaccination (\*)** |  |  |  |  |
| Mean [Range] | 31 [23-65] | 55 [24-81] | 42 [32-52] | 32 [23-81] |
| **Sex at Birth (ns)** |  |  |  |  |
| Female | 58 (70%) | 8 (53%) | 2 (100%) | 68 (68%) |
| Male | 25 (30%) | 7 (47%) | 0 (0%) | 32 (32%) |
| **Race (\*\*)** |  |  |  |  |
| Asian | 20 (24%) | 1 (7%) | 0 (0%) | 21 (21%) |
| Black or African American | 7 (8%) | 0 (0%) | 0 (0%) | 7 (7%) |
| Middle Eastern or North African | 1 (1%) | 1 (7%) | 0 (0%) | 2 (2%) |
| White | 39 (47%) | 7 (47%) | 1 (50%) | 47 (47%) |
| More Than One Race | 9 (11%) | 2 (13%) | 0 (0%) | 11 (11%) |
| Other | 7 (8%) | 1 (7%) | 1 (50%) | 9 (9%) |
| Unknown | 0 (0%) | 3 (20%) | 0 (0%) | 3 (3%) |
| **Vaccine Manufacturer** |  |  |  |  |
| Abbott | 0 (0%) | 1 (7%) | 0 (0%) | 1 (1%) |
| GSK | 0 (0%) | 6 (40%) | 0 (0%) | 6 (6%) |
| Sanofi | 0 (0%) | 5 (33%) | 2 (100%) | 7 (7%) |
| Seqirus | 83 (100%) | 3 (20%) | 0 (0%) | 86 (86%) |
| **Vaccine Trade Name** |  |  |  |  |
| Afluria | 0 (0%) | 2 (13%) | 0 (0%) | 2 (2%) |
| Fluad | 0 (0%) | 1 (7%) | 0 (0%) | 1 (1%) |
| Fluarix | 0 (0%) | 6 (40%) | 0 (0%) | 6 (6%) |
| Flublok | 0 (0%) | 0 (0%) | 2 (100%) | 2 (2%) |
| Flucelvax | 83 (100%) | 0 (0%) | 0 (0%) | 83 (83%) |
| Fluzone | 0 (0%) | 3 (20%) | 0 (0%) | 3 (3%) |
| Fluzone High Dose | 0 (0%) | 2 (13%) | 0 (0%) | 2 (2%) |
| Influvac | 0 (0%) | 1 (7%) | 0 (0%) | 1 (1%) |
| **Simultaneous COVID-19 Vaccine?** |  |  |  |  |
| Yes | 15 (18%) | 6 (40%) | 0 (0%) | 21 (21%) |
| No | 68 (82%) | 9 (60%) | 2 (100%) | 79 (79%) |
| **Previous Vaccine Frequency** |  |  |  |  |
| Every Year | 74 (89%) | 14 (93%) | 1 (50%) | 89 (89%) |
| Intermittently | 8 (10%) | 1 (7%) | 1 (50%) | 10 (10%) |
| Never | 1 (1%) | 0 (0%) | 0 (0%) | 1 (1%) |
| **Comorbidities?** |  |  |  |  |
| Yes | 5 (6%) | 4 (27%) | 1 (50%) | 10 (10%) |
| No | 56 (67%) | 9 (60%) | 1 (50%) | 66 (66%) |
| Not Reported | 22 (27%) | 2 (13%) | 0 (0%) | 24 (24%) |

### Characterization of the antigenic mismatch

We measured HI titers using sequence-verified viral stocks of four H3N2 isolates representative of the vaccine strain (A/New York City/PX12775/2024, [EPI_ISL_19695815]; vaccine-like (VL) strain), the canonical subclade K (A/New York City/PX23710/2025, [EPI_ISL_20330074] clade K root (K)) as well as two subclade K strains with two or three additional, distinct mutations in HA (A/New York City/PX23641/2025, same as clade K root plus HA1: R329K, HA2: A43S (KS) [EPI_ISL_20330072]; A/New York City/PV307069/2025, same as clade K root plus G62E, S145R, R208K (ERK), [EPI_ISL_20315134], Supplemental Table 1 for additional information).

Prior to vaccination, HI titers for the vaccine-like isolate and two of the subclade K variants were comparable (GMT: 15-16) with a third of the pre-vaccine sera (31-37%) being non-reactive (Fig. 1B). Recognition of the third subclade K isolate, A/New York City/PV307069/2025 (ERK), was more modest, with over half of the sera (53%) failing to inhibit hemagglutination of guinea pig red blood cells (Fig. 1B). Influenza vaccination increased the HI titers by 2-fold (p<0.05) for the subclade K viruses and by 3-fold (p<0.0001) for the vaccine strain (Fig. 1C). Importantly, half of the participants with non-reactive titers prior to vaccination mounted immune responses to one or more of the viruses tested after vaccination (HI titer ≥10). Across all four viruses, cell-based and recombinant vaccines showed statistically significant pre to post vaccination rises with two-to-three-fold increases in geometric mean titers (GMT).

Historically, an HI titer of 1:40 has been associated with a 50% reduction in risk of influenza. The proportion of participants with HI titers ≥40 ranged between 16%-32% prior to vaccination and increased to 39%–65% after vaccination, with the lowest responses corresponding to the most mutated of the subclade K variants (A/New York City/PV307069/2025 (ERK)) and the strongest responses to A/New York City/PX12775/2024 (VL), the vaccine-like virus (Fig. 1D). Egg-based vaccines (n=15) induced a similar fold increase for A/New York City/PX23710/2025 (K) (2-fold) but more modest increases for A/New York City/PX12775/2024 (VL), A/New York City/PX23641/2025 (KS) and A/New York City/PV307069/2025 (ERK). The proportion of participants with HI titers ≥40 ranged between 53% and 67% post-immunization with inactivated egg-based vaccines except again for the most mutated variant A/New York City/PV307069/2025 (ERK) (HI ≥40: 27%). All age groups showed increases in HI titers post-vaccination, but the magnitude varied by age group and virus (Fig. 1F). While fold-rises decreased with age, potentially due to higher baseline immunity or immunosenescence, all age groups demonstrated vaccine responsiveness.

Participants with higher baseline HI titers tended to have smaller fold-rises post-vaccination, consistent with the concept of an antibody ceiling where pre-existing immunity limits the magnitude of boost responses^8–10^. In contrast, we observed higher than average fold increases with similar rates of seroconversion for participants with HI titers below the limit of detection prior to vaccination: vaccine-like virus (4-fold, 47% seroconversion), A/New York City/PX23710/2025 (K) (5-fold, 51% seroconversion), A/New York City/PX23641/2025 (KS) (4-fold, 45% seroconversion) and A/New York City/PV307069/2025 (ERK) (7-fold, 42% seroconversion). Sera from 8% of participants remained HI non-reactive for all four viruses both before and after vaccination.

Neutralization titers were measured on hCK cells^11^ for the vaccine-like A/New York City/PX12775/2024 (VL) and the subclade K root representative A/New York City/PX23710/2025 (K) viral isolates. In agreement with the HI titers, we noted pre-existing neutralization titers to both vaccine-matched and antigenically distinct subclade K virus (GMT 14 and 12) with 70% and 59% seroreactivity, respectively (Fig. 2A). Upon vaccination, titers for both viruses increased two-fold (p<0.01) but the percent seropositivity did not change substantially (Fig. 2B) relative to the pre-vaccine levels. HI and neutralization titers showed strong correlation for both the vaccine strain (Spearman r=0.42, p<.0001, Fig. 2C) and subclade K variant (r=0.51, p<0.001, Fig. 2D), confirming HI as a reliable surrogate for neutralizing activity against these viruses.

**Figure 2:**
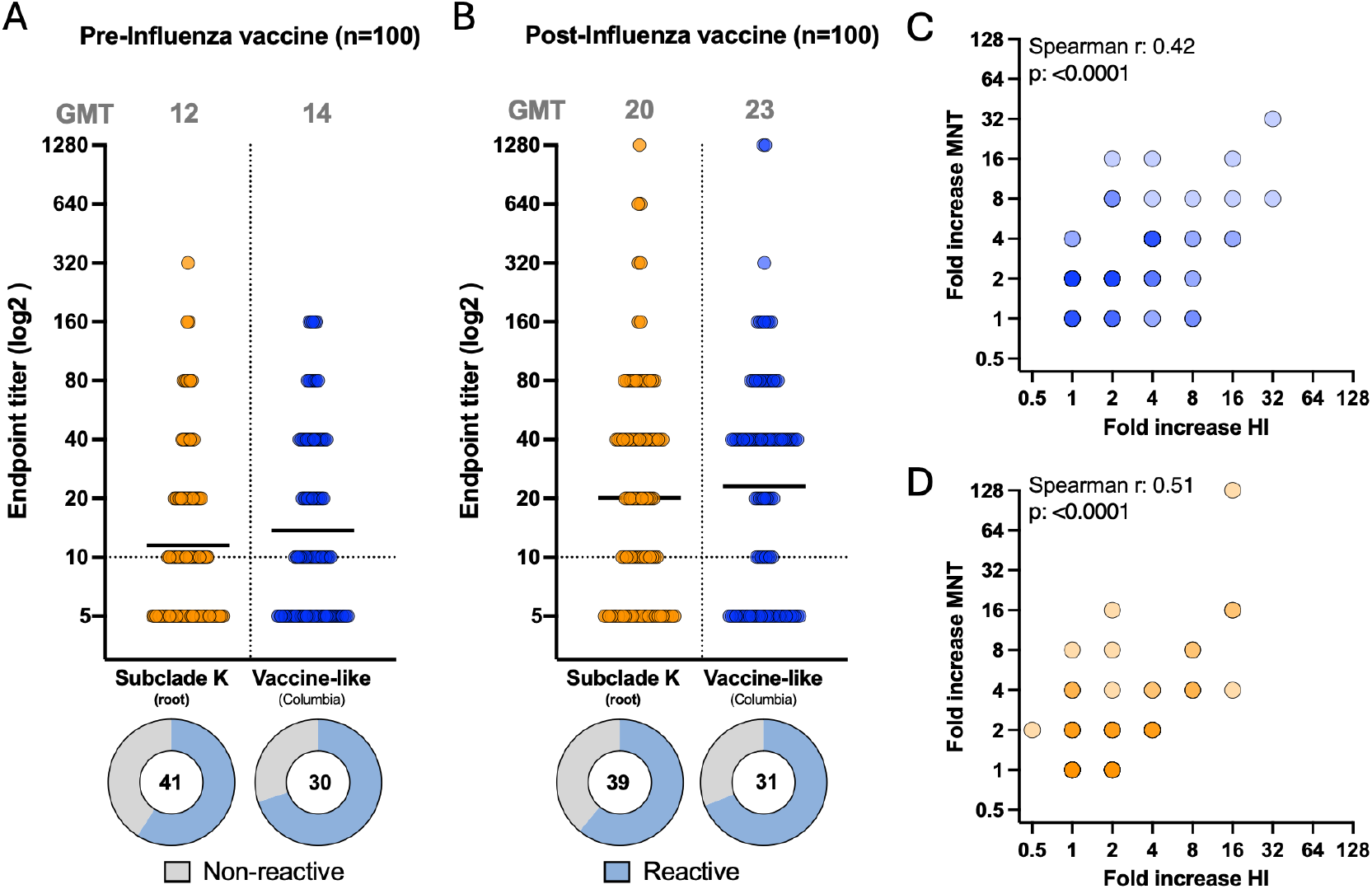
Microneutralization titers against H3N2 subclade K and vaccine like isolates before and after seasonal immunization. **A/B**: The pre- and post-influenza vaccine microneutralization titers for 200 sera are shown for the two viruses tested. The donuts illustrate the proportion of reactive (endpoint titer: ≥10) and non-reactive sera. with the number of sera with values below the limit of detection being shown for each virus and time point. **C/D**: Comparisons of the fold increase in HI titers compared to fold increase in microneutralization titers for the vaccine-like strain (blue) and the subclade K (root, yellow) viral variant are shown.

## Conclusions

In this study, we assessed serologic responses to subclade K in a metropolitan adult cohort with substantial prior seasonal influenza vaccination. Although H3N2 subclade K variants caused record-high infection rates in the Northern Hemisphere, our findings demonstrate that the 2025/2026 seasonal influenza vaccine formulation increased HI and neutralization titers against both the vaccine strain and multiple subclade K viruses.

These observations are consistent with the vaccine effectiveness assessments in the United Kingdom^4^, China^12^, Canada^13^ and the European Union^6^ which indicated that vaccination remained effective against clinical disease caused by influenza A (H3N2) viruses. There is a notable discrepancy between the modest decrease in antibody titers for subclade K variants observed in sera from adults living in a large US metropolitan area and the substantial drop in HI titers for subclade K variants observed when using ferret sera raised against vaccine strains^4^. Similar findings have, however, been reported by other groups using human biospecimen^14^. Thus, there is an urgent need to use human sera for continued antigenic monitoring to guide strain selection in a manner that considers the contribution of pre-existing immunity in adult populations.

## Material and Methods

### Human sera

We selected 200 sera from 100 adult study participants of two observational longitudinal institutional review board (IRB)-approved cohort studies conducted in New York City (STUDY-16-01199 | STUDY-16-01215). All participants provided written informed consent prior to data/sample collection. Sera were collected in the fall of 2025 before and after study participants were immunized with the 2025/2026 formula of the seasonal influenza vaccine per their standard of care. Sera was stored at −80°C until use. 83% of the participants elected to receive the cell-based (Flucelvax, Seqirus), 15% elected egg-based (e.g., Fluarix, GlaxoSmithKline; Fluzone, Sanofi) and 2% elected recombinant (Flublok, Sanofi) 2025/2026 influenza vaccines. The prior seasonal influenza vaccine frequency varied among participants, with 89% of the participants having been immunized annually and 10% reporting intermittent vaccination. One participant reported receiving the seasonal influenza vaccine for the very first time. Detailed demographics and metadata for each group are provided in Table 1.

### Influenza A H3N2 virus isolates

The Mount Sinai Pathogen Surveillance Program leverages residual clinical biospecimen that tested positive for influenza viruses to track the spread and evolution of disease-causing influenza viruses. The study protocol was IRB-approved (STUDY-13-00981). Nasopharyngeal swabs were selected for culture based on the HA sequences. Briefly, 200ul of viral transport media was incubated with hCK cells^11^ at 37°C and culture supernatants were harvested once cytopathic effects were visible. Sequence-verified stocks representing the vaccine strain (A/New York City/PX12775/2024, [EPI_ISL_19695815]) and three subclade K isolates (A/New York City/PX23710/2025 [EPI_ISL_20330074], similar to the root of the subclade K; A/New York City/PX23641/2025, clade K plus HA1:R329K, HA2:A43S [EPI_ISL_20330072]; A/New York City/PV307069/2025, same as clade K root plus G62E, S145R, R208K, [EPI_ISL_20315134]) were used to measure HI and neutralization titers.

### Hemagglutination inhibition assays (HI)

The 200 sera were treated overnight with receptor-destroying enzyme (RDE) from Vibrio cholerae (Denka Seiken, Tokyo, Japan) at 37 °C. Following incubation, 2.5% sodium citrate was added, and the mixture was heat-inactivated at 56 °C for 60 min. RDE-treated sera were then diluted to a starting concentration of 1:10 with PBS (1X). Subsequently, 50 μL of each treated serum sample was subjected to two-fold serial dilution in PBS (1X) in V-bottom 96-well microplates (NUNC), reaching final dilutions up to 1:2560. An equal volume (50 μL) of virus standardized to four hemagglutinating units was added to each well, and the plates were incubated on a shaker for 30 min at room temperature. Following incubation, 50 μL of 0.5% guinea pig red blood cells (RBCs; cat. no. IGPRBC10ML, Innovative Research) prepared in PBS (1X) were added to the serum-virus mixture, and plates were incubated at 4 °C for 60–90 min. HI titers were recorded as the reciprocal of the highest serum dilution that completely inhibited hemagglutination by four units of influenza virus.

### Microneutralization (MNT) assay

Humanized Madin-Darby canine kidney (hMDCK or hCk) cell line expressing high levels of human influenza virus receptors and low levels of avian virus receptors cells adapted to human influenza A/H3N2 strains^11^ were seeded in 96-well cell-culture treated plates (Corning) at density of 2 × 10^5^ cells/mL (100 μL/well) and incubated at 37°C with 5% CO_2_ overnight. The following day, RDE-treated human sera were initially diluted 1:10 and serially diluted 2-fold across the plates in infection media consisting of 1x minimum essential media (MEM) (Gibco), 100 U/mL penicillin and 100 μg/mL streptomycin (Gibco), 10 mM 4-(2-hydroxyethyl)-1-piperazineethanesulfonic acid (HEPES) (Gibco), 2 mM L-glutamine (Gibco), 3.2% NaHCO_3_ (Sigma-Aldrich), and 1.2% bovine serum albumin (BSA) (MP Biomedicals) supplemented with 1 μg/mL N-tosyl-L-phenylalanine chloromethyl ketone (TPCK) - treated trypsin (Sigma-Aldrich). Next, 60 μL of 2000 × 50% tissue culture infectious dose (TCID_50_) per mL of virus prepared in infection medium and 60 μL of serially diluted sera were incubated on a shaker at RT for 1 h to allow antibodies to bind to virions. Before the end of the incubation, hCK cells were washed with 220 μL of PBS (1X) and incubated with 100 μL of the incubated serum-virus mixture at 37 °C with 5% CO_2_ for 1 h to allow for attachment of virions to the cells. Afterwards, the virus inoculum was carefully aspirated, hCK cells were washed with PBS (1X), and 50 μl of serially diluted sera was transferred to corresponding wells on hCK microplates. All the wells were supplemented with 50 μl of infection media, and cell microplates were incubated for 48h at 37°C with 5% CO_2_. Viral neutralization was assessed using a conventional hemagglutination assay as described above. Briefly, 50 μL of cell culture supernatant was mixed with 50 μL of 0.5% guinea pig RBCs (cat. no. IGPRBC10ML, Innovative Research) diluted in PBS (1X) in V-bottom 96-well plates (NUNC). Following incubation at 4 °C for 60-90 min, neutralization titers were defined as the reciprocal of the highest serum dilution that completely inhibited hemagglutination.

## Statistics

All statistical analyses in the full cohort were conducted with GraphPad Prism (version 10.3). Normality was tested using a Shapiro-Wilk test. Differences between the four groups were analyzed with one-way ANOVA non-parametric test using Kruskal–Wallis test with Dunn’s correction. Correlations between HI and microneutralization titers were analyzed using Spearman’s rank test. Significance was considered with p-values equal or less than 0.05 (*), ≥0.01 (**), ≥0.001 (***), ≥0.0001 (****). Only significant differences (p < 0.05) between GMTs of pre- and post-vaccination are shown on plots.

## Data Availability

All data produced in the present study are available upon reasonable request to the authors

## Acknowledgments

We thank all the study participants for their generous support of our research programs. This work was funded through the NIAID Centers for Excellence in Influenza Research and Response (75N93021C00014), the Viral Immunity and Vaccination (*VIVA*) Human Immunology Project Consortium (*HIPC*) (NIAID U19 AI168631) and through institutional support from the Mount Sinai Center for Vaccine Research and Pandemic Preparedness. Work at the Medical University of Vienna was funded by institutional funds.

---

**Supplementary Table 1:** Detailed information regarding the four H3N2 isolates.

| Isolate_Name | Subtype | Clade |  | Isolate_Id | Month of collection |  |
| --- | --- | --- | --- | --- | --- | --- |
| A/New York/PX12775/2024 | A / H3N2 | 3C.2a1b.2a.2a.3a.1 |  | EPI_ISL_19695815 | Dec-24 |  |
| A/New York/PX23710/2025 | A / H3N2 | 3C.2a1b.2a.2a.3a.1 / K |  | EPI_ISL_20330074 | Oct-25 |  |
| A/New York/PX23641/2025 | A / H3N2 | 3C.2a1b.2a.2a.3a.1 / K |  | EPI_ISL_20330072 | Oct-25 |  |
| A/New York/PV307069/2025 | A / H3N2 | 3C.2a1b.2a.2a.3a.1 / K |  | EPI_ISL_20315134 | Nov-25 |  |
| Isolate_Name | Subtype | PB2 Segment_Id |  | PB1 Segment_Id | PA Segment_Id | MP Segment_Id |
| A/New York/PX12775/2024 | A / H3N2 | EPI3862970 \| 71347 \| PX12775 \| PB2 |  | EPI3862968 \| 71347 \| PX12775 \| PB1 | EPI3862965 \| 71347 \| PX12775 \| PA | EPI3862971 \| 71347 \| PX12775 \| MP |
| A/New York/PX23710/2025 | A / H3N2 | EPI5011343 \| 80573 \| PX23710 \| PB2 |  | EPI5011341 \| 80573 \| PX23710 \| PB1 | EPI5011338 \| 80573 \| PX23710 \| PA | EPI5011344 \| 80573 \| PX23710 \| MP |
| A/New York/PX23641/2025 | A / H3N2 | EPI5011327 \| 80571 \| PX23641 \| PB2 |  | EPI5011325 \| 80571 \| PX23641 \| PB1 | EPI5011322 \| 80571 \| PX23641 \| PA | EPI5011328 \| 80571 \| PX23641 \| MP |
| A/New York/PV307069/2025 | A / H3N2 | EPI4971339 \| 81477 \| PV307069 \| PB |  | EPI4971337 \| 81477 \| PV307069 \| PB1 | EPI4971334 \| 81477 \| PV307069 \| PA | EPI4971340 \| 81477 \| PV307069 \| MP |
| Isolate_Name | Subtype | HA Segment_Id |  | NP Segment_Id | NA Segment_Id | NS Segment_Id |
| A/New York/PX12775/2024 | A / H3N2 | EPI3862966 \| 71347 \| PX12775 \| HA |  | EPI3862967 \| 71347 \| PX12775 \| NP | EPI3862964 \| 71347 \| PX12775 \| NA | EPI3862969 \| 71347 \| PX12775 \| NS |
| A/New York/PX23710/2025 | A / H3N2 | EPI5011339 \| 80573 \| PX23710 \| HA |  | EPI5011340 \| 80573 \| PX23710 \| NP | EPI5011337 \| 80573 \| PX23710 \| NA | EPI5011342 \| 80573 \| PX23710 \| NS |
| A/New York/PX23641/2025 | A / H3N2 | EPI5011323 \| 80571 \| PX23641 \| HA |  | EPI5011324 \| 80571 \| PX23641 \| NP | EPI5011321 \| 80571 \| PX23641 \| NA | EPI5011326 \| 80571 \| PX23641 \| NS |
| A/New York/PV307069/2025 | A / H3N2 | EPI4971335 \| 81477 \| PV307069 \| HA |  | EPI4971336 \| 81477 \| PV307069 \| NP | EPI4971333 \| 81477 \| PV307069 \| NA | EPI4971338 \| 81477 \| PV307069 \| NS |
| Isolate_Name | Subtype | Genbank Accessions |  |  |  |  |
| A/New York/PX12775/2024 | A / H3N2 | NA:PV063985,PB2:PV063986,PA:PV063987,NS:PV063988,PB1:PV063989,NP:PV063990,HA:PV063991,MP:PV06 |  |  |  |  |
| A/New York/PX23710/2025 | A / H3N2 | PB1:PX903194,HA:PX903195,NA:PX903196,MP:PX903197,PB2:PX903198,NS:PX903199,NP:PX903200,PA:PX90 |  |  |  |  |
| A/New York/PX23641/2025 | A / H3N2 | MP:PX903946,PB1:PX903947,NS:PX903948,HA:PX903949,PB2:PX903950,PA:PX903951,NP:PX903952,NA:PX90 |  |  |  |  |
| A/New York/PV307069/2025 | A / H3N2 | PB1:PX902587,MP:PX902588,NA:PX902589,NP:PX902590,PA:PX902591,HA:PX902592,NS:PX902593,PB2:PX90 |  |  |  |  |

